# Association between calcium and vitamin D in geriatric patients hospitalized for Covid-19: Results from the GERIA-COVID study

**DOI:** 10.1101/2023.02.07.23285612

**Authors:** Alexis Bourgeais, Jennifer Gautier, Guillaume Sacco, Cédric Annweiler, Marine Asfar, the GERIA-COVID study group

## Abstract

The deficiency of 25OH vitamin D (25[OH]D) is common in the older population. It physiologically triggers secondary hyperparathyroidism resulting in normal circulating calcium levels. Adjusted calcium (CaA) is estimated by the PAYNE method and several studies report a misclassification of calcium status by corrected calcium compared to ionized calcium (CaI) in older patients. Hypocalcemia is common in older COVID-19 patients. Blunted secondary hyperparathyroidism explain this high prevalence of hypocalcemia in COVID-19. However, no studies have focused on patients older than 75 years despite the high mortality rate in this population. In the present study, the association between the different types of calcium (CaI, CaA, and total calcium [CaT]) and 25(OH)D deficiency (below 50 nmol/L) was investigated. The study of the correlation between each type of calcium was performed secondarily. Observational monocentric study focused on the GERIA-COVID database during the second wave of COVID-19 in France from October 2020 to March 2021. COVID-19 was diagnosed with RT-PCR and/or chest CT-scan. A population of 181 older COVID-19 patients (86.4 years ± 5.7) was analyzed. Sixty-three patients (34.8%) were deficient in 25(OH)D. The prevalence of total and ionized hypocalcemia was 44.1% and 39.2%, respectively. A negative association was reported in linear regression between 25(OH)D deficiency and CaA (β =-0.052 [- 0.093; -0.010], p = 0.015) as well as with CaT (β = -0.05 [-0.09; -0.01], p =0.034) in the multivariate model. No association was found between vitamin D deficiency and CaI. In the multivariate models, there was no association between each type of calcium and PTH. CaI was correlated with CaT (r = 0.39, p < 0.001) and with CaA (r = 0.15, p = 0.043). Secondary hyperparathyroidism was not activated in the context of COVID-19 in this study. After reviewing the literature, this appears to be the first study in older patients to expose such results.

## INTRODUCTION

Vitamin D refers to a steroid hormone. It is ingested as ergosterol (D2) or cholecalciferol (D3), or supplied as D3 from the conversion of 7-dehydrocholesterol in the skin by ultraviolet B radiation. D2 or D3 are synthesized in the liver to 25-hydroxyvitamin D [25(OH)D], the major metabolite of vitamin D in circulation. The 25(OH)D is hydroxylated to 1,25-dihydroxyvitamin D [1,25(OH)D3] which is the metabolically active form of vitamin D (1). According to different studies, 25(OH)D deficiency is a relatively common disorder in older population with a prevalence between 40 and 100% (2).

Both 1,25(OH)D3 and Parathormone (PTH) regulate calcium absorption. When 1,25(OH)D3 levels are low, secondary hyperparathyroidism increases renal conversion of 25(OH)D and thus maintains normal plasma levels of 1,25(OH)D3 until the 1,25(OH)D3 deficiency is severe enough to reduce the level of this metabolite (1–3). Fibroblast growth factor-23, which is a protein, is also involved in this complex metabolism by stimulating the conversion to 1,25(OH)D3 (4).

Most of the organism’s calcium is chelated to phosphorus in the bones (99.9 %). Circulating calcium (0.07%) is separated into two elements: ionized calcium and calcium bound to protein. Ionized calcium (CaI) (40-50%) is considered the active element, and calcium bound to albumin or other proteins represents 50-60% of the circulating calcium (5).

Because CaI is expensive and difficult to measure, formulas are used in clinical practice to estimate calcium concentration. The best known and most widely used formula is Payne’s, defined as follows: (*0*.*025 x (40 – albumin [g/L]) + total calcium [CaT, mmol/L]) = adjusted calcium (CaA, mmol/L)* (6,7).

Nevertheless, this formula does not consider the non-protein parameters involved in calcium regulation and could misclassify the calcium status of older patients (8–10).

Hypocalcemia is frequently associated with COVID-19 (11,12); the mechanism of these imbalances in calcium metabolism is not fully understood. Furthermore, none of the studies conducted to investigate the relationship between COVID-19, 25(OH)D and hypocalcemia have focused on the older population (12,13). Paradoxically, the mortality rate of this disease in older patients is high and increases with age (14). In COVID-19, it is clinically important to perform a detailed assessment of serum calcium and to have an increasingly accurate understanding of its mechanisms in the older population.

In this context, we aimed to evaluate the association between the different types of calcium (CaT, CaA, CaI) and deficiency in 25(OH)D in the older patients hospitalized for COVID-19. Secondary objectives were to evaluate the correlation between CaI and the two others type of calcium (adjusted and total).

## METHODS

An observational retrospective study based on the GERIA-COVID database was performed. GERIA-COVID study, which was described elsewhere (15), was carried out in the geriatric acute care unit dedicated to COVID-19 patients in the University Hospital of Angers, France. Data analyzed in our study focused on the second wave (October 2020 - March 2021) of COVID-19 pandemic.

### Study population

The inclusion criteria in the present study were: 1) Patients aged 75 years and over hospitalized in the geriatric acute care unit of Angers University Hospital, France, between October 2020 and March 2021; 2) Data available on ionized calcium and total calcium concentrations; 3) COVID-19 diagnosed with RT-PCR and/or chest CT-scan; 4) No objection from the patient and/or relatives to the use of anonymized clinical and biological data for research purpose. (Figure 1)

**Figure 1:**
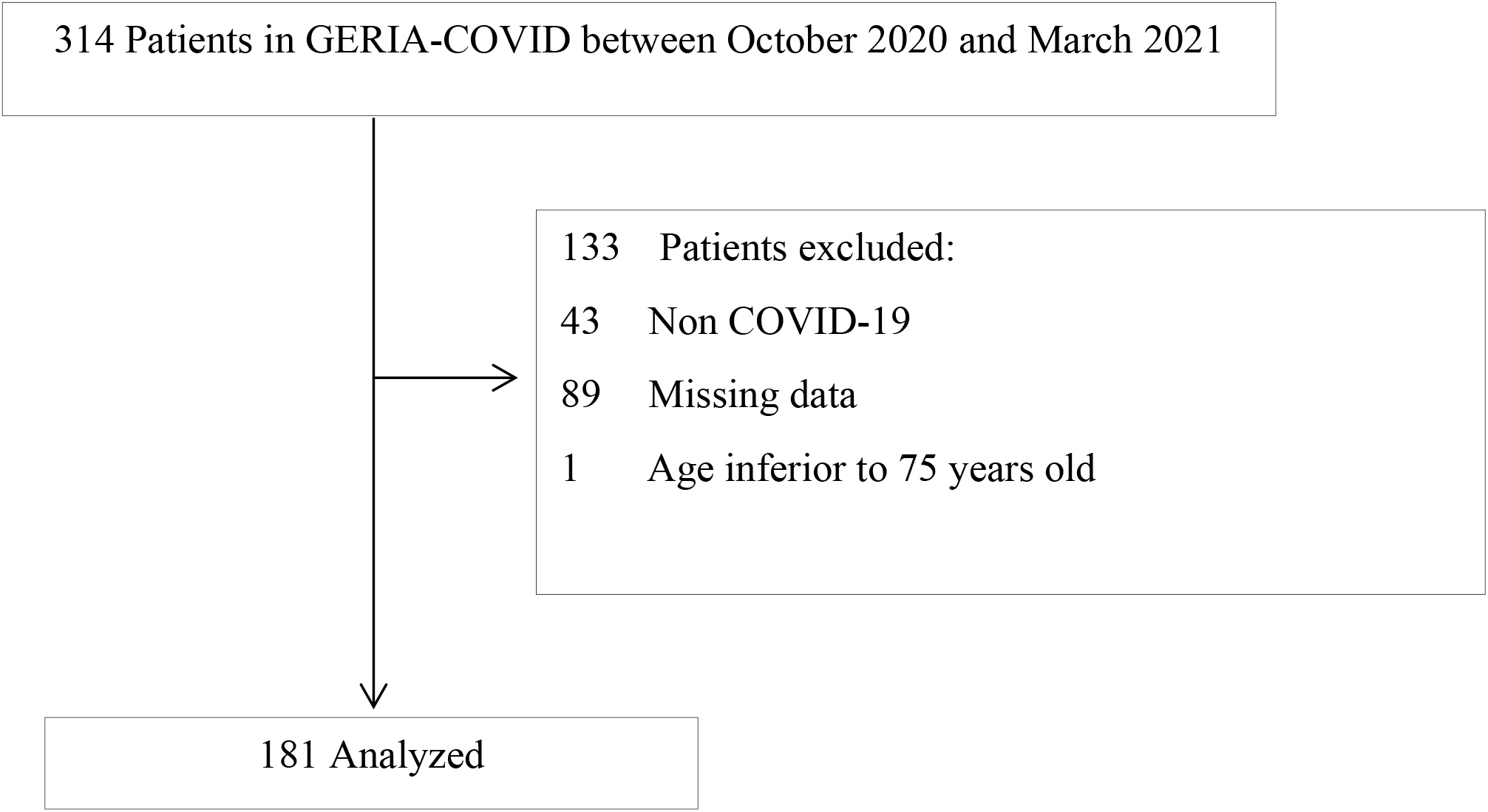
Flow chart.

### Biological data and COVID-19 diagnostic

All biological samples were analyzed at the Biology department of the University Hospital Angers. COVID-19 was diagnosed with RT-PCR and/or chest CT-scan. Ionized calcium was collected with a blood gas syringe and analyzed by blood gas machine from the first of October 2020 to the 29^th^ of November then by direct potentiometry (ABL 90 Flex plus or ABL 800) from the 30^th^ of November 2020 to the 31^st^ of March 2021. This measurement was systematically adjusted to pH. An immunoassay method (Automate LiaisonXL, DiaSorin) was used to measure PTH and 25(OH)D.

C-Reactive Protein (CRP) and albumin were measured by immunoturbidimetric method (Advia Chemistry XPT, Siemens). Albumin was measured on the same blood sample than calcium. Colorimetric method was used to measure total calcemia. Glomerular filtration rate has been estimated with Modification of Diet in Renal Disease (MDRD). The Payne’s method was used to correct the calcemia: *(0*.*025 x (40 – albumin [g/L]) + total calcium [CaT, mmol/L]) = adjusted calcium (CaA, mmol/L)*.

### Statistical analysis

A descriptive analysis of the participant’s characteristics was firstly performed using effectives and percentages for qualitative variables and means ± standard deviations or medians [inter-quartile range (IQR)] for quantitative variables, as appropriate. Univariate linear regressions were performed to measure the association between 25(OH)D deficiency (ie < 50 nmol/L) and each type of calcium (CaT, CaA, CaI). Then, multivariable models were completed considering these covariates: age, sex, PTH, creatinin, severe denutrition (defined by an albuminemia inferior to 30 g/L). Spearman’s correlation test was used to measure the correlation between the three different types of calcemia and 25(OH)D. Association or correlation was assessed if p <0.05. Analyses were performed using SAS® version 9.4 (SAS Institute Inc.).

### Ethics

The study was conducted in accordance with the ethical standards set forth in the Helsinki Declaration (1983). No participant or relatives objected to the use of anonymized clinical and biological data for research purposes. Ethics approval was obtained from the Ethics Board of the University Hospital of Angers, France (2022-102).

## RESULTS

### Clinical characteristics of the population

A total of 181 patients were included in this study, including 95 women (50.8%), mean age was 86.4 ± 5.7 years. Sixty-three patients suffered from severe denutrition among the participants (34.8 %) and 63 participants (34.8%) were deficient in 25(OH)D. The medians of CaT, CaA and CaI were 2.22 [2.14-2.32], 2.42 [2.34 – 2.50] and 1.19 [1.15-1.23] mmol/L, respectively. The means of PTH and creatinin were 29.7 [18.8 – 43] pg/mL and 72 [56 – 104] µmol/L, respectively (Table I).

**Table I:** Participant’s characteristics (N = 181)

|  | Total<br>(N = 181) |
| --- | --- |
| **Demographic characteristics** |  |
| Age, years, mean ± SD | 86.4±5.7 |
| Female gender, n (%) | 92 (50.8) |
| Severe undernutrition, n (%) * | 63 (34.8) |
| **Biology** |  |
| PTH, pg/mL, med [IQR] | 29.7 [18.8-43.0] |
| Creatinin, μmol/L, med [IQR] | 72 [56-104] |
| Total calcemia, mmol/L, med [IQR] | 2.22 [2.14-2.32] |
| Corrected calcemia, mmol/L, med [IQR] | 2.42 [2.34-2.50] |
| Ionized calcemia, mmol/L, med [IQR] | 1.19 [1.15-1.23] |
| Total hypocalcemia (<2.2 mmol/L), n (%) | 80 (44.1) |
| Adjusted hypocalcemia (<2.2 mmol/L), n (%) | 9 (5.0) |
| Ionized hypocalcemia (<1.18 mmol/L), n (%) | 71 (39.2) |
| 25(OH)D, nmol/mL, med [IQR] | 66 [36-86] |
| 25(OH)D < 50 nmol/mL, n (%) | 63 (34.8) |

SD: Standard Deviation; IQR: Interquartile Ranges; \* Albuminemia < 30 g/L; med : median

### Association between calcium and deficiency in 25(OH)D

This study performed linear regressions between each type of calcium and deficiency in 25(OH)D. In Table II, a significant negative association was found between CaA and deficiency in 25(OH)D whether analyzed in univariate (β = - 0.054 [-0.100; -0.007], p < 0.021) or multivariate model (β = - 0.052 [-0.093; -0.010], p < 0.015). Age and severe denutrition were also positively associated with CaA in univariate model (respectively, p < 0.012 and p < 0.001) and remained significant in the multivariate model (respectively, p < 0.019 and p < 0.001). PTH was not significantly associated with CaA in the univariate model, nor in the multivariate model.

**Table II.** Linear regressions between adjusted calcium (CaA) and deficiency in 25(OH)D (< 50 nmol/L) (N = 181)

|  | Univariate models |  | Fully adjusted model |  |
| --- | --- | --- | --- | --- |
| | $\beta$ [95% CI] | P-value | $\beta$ [95% CI] | P-value |
| 25(OH)D < 50 nmol/L | -0.054 [-0.100 ; -0.007] | **0.021** | -0.052 [-0.093 ; -0.010] | **0.015** |
| Age | 0.005 [0.001 ; 0.009] | **0.012** | 0.004 [0.001 ; 0.008] | **0.019** |
| Female gender | 0.04 [0.00 ; 0.08] | 0.063 | 0.04 [0.00 ; 0.08] | 0.059 |
| Severe undernutrition † | 0.14 [0.10 ; 0.18] | **<0.001** | 0.14 [0.10 ; 0.18] | **<0.001** |
| PTH, for 100 pg/mL | -0.05 [-0.12 ; 0.02] | 0.142 | -0.02 [-0.10 ; 0.06] | 0.625 |
| Creatinin, for 100 μmol/L | -0.02 [-0.05 ; 0.02] | 0.316 | -0.02 [-0.06 ; 0.01] | 0.197 |

†Albuminemia < 30 g/L

In Table III, univariate models showed no significant association between CaT and deficiency in 25(OH)D. Nevertheless after adjustment on age, gender, severe denutrition, PTH and creatinin, we found a significant negative association (β = - 0.05 [-0.09; -0.01], p = 0.034). The other significantly associated variables with CaT were creatinin (p = 0.005), gender (p < 0.001) and severe denutrition (p = 0.002). These covariables remained associated in the multivariate model. PTH was not significantly associated with CaT in the univariate model, nor in the multivariate model.

**Table III.** Linear regressions between total calcium (CaT) and deficiency in 25(OH)D (< 50 nmol/L) (N = 181)

|  | Univariate models |  | Fully adjusted model |  |
| --- | --- | --- | --- | --- |
| | $\beta$ [95% CI] | P-value | $\beta$ [95% CI] | P-value |
| 25(OH)D < 50 nmol/L | -0.04 [-0.08 ; 0.01] | 0.092 | -0.05 [-0.09 ; -0.01] | **0.034** |
| Age | 0.003 [-0.001 ; 0.006] | 0.172 | 0.003 [-0.001 ; 0.006] | 0.118 |
| Female gender | 0.07 [0.03 ; 0.11] | **<0.001** | 0.045 [0.005 ; 0.086] | **0.029** |
| Severe undernutrition † | -0.07 [-0.11 ; -0.03] | **0.002** | -0.06 [-0.10 ; -0.02] | **0.004** |
| PTH, for 100 pg/mL | -0.03 [-0.10 ; 0.03] | 0.332 | 0.01 [-0.07 ; 0.09] | 0.723 |
| Creatinin, for 100 μmol/L | -0.04 [-0.07 ; -0.01] | **0.005** | -0.04 [-0.077 ; -0.004] | **0.031** |

†Albuminemia < 30 g/L

In Table IV, we found no association between CaI and deficiency in 25(OH)D in our population no matter univariate or multivariate model. PTH was not significantly associated with CaI in the univariate model nor in the multivariate model.

**Table IV.** Linear regressions between ionized calcium (CaI) and deficiency in 25(OH)D (< 50 nmol/L) (N = 181)

|  | Univariate models |  | Fully adjusted model |  |
| --- | --- | --- | --- | --- |
| | $\beta$ [95% CI] | P-value | $\beta$ [95% CI] | P-value |
| 25(OH)D < 50 nmol/L | -0.02 [-0.04 ; 0.01] | 0.129 | -0.02 [-0.05 ; 0.01] | 0.080 |
| Age | 0.002 [-0.001 ; 0.004] | 0.095 | 0.002 [0.001 ; 0.004] | **0.039** |
| Female gender | 0.01 [-0.01 ; 0.03] | 0.339 | 0.00 [-0.02 ; 0.02] | 0.949 |
| Severe undernutrition † | -0.02 [-0.4 ; 0.01] | 0.129 | -0.02 [-0.04 ; 0.01] | 0.147 |
| PTH, for 100 pg/mL | -0.02 [-0.06 ; 0.02] | 0.261 | 0.00 [-0.05 ; 0.05] | 0.995 |
| Creatinin, for 100 μmol/L | -0.018 [-0.034 ; -0.002] | **0.031** | -0.02 [-0.04 ; 0.01] | 0.054 |

†Albuminemia < 30 g/L

### Correlation between each type of calcium

No significant correlation with Spearman’s test was observed between 25(OH)D and the different types of calcemia. This study showed a moderate correlation between total calcium and CaA (r = 0.48, p < 0.001). CaI and CaT were also moderately correlated (r = 0.39, p < 0.001). There was a slight correlation between CaA and CaI (r = 0.15, p = 0.043).

## DISCUSSION

In this study, a negative association was found between CaT, CaA and 25(OH)D deficiency in older hospitalized patients for COVID-19. Patients with 25(OH)D deficiency had significantly lower level of calcium (total and adjusted) than those without deficiency. We demonstrated no association between CaI and deficiency in 25(OH)D. In addition, PTH was not shown to be significantly associated with any of the three types of calcium levels in the multivariate model.

Hypocalcemia is frequent in the covid context: 35 to 59 % of the patients have low level of calcium, which is in agreement with our report of ionized and total calcium analysis described in table I (16–18). Di Filippo et al. also did not describe an association between 25(OH)D deficiency and ionized calcium, but showed an association between total calcium and 25(OH)D deficiency. They described the absence of secondary hyperparathyroidism in younger COVID-19 patients; the median age being 55.9 [48.5-64.5] years in their study. The absence of calcium adjustment is a limitation in their study (13).

Hypothesis to explain our results could be the suppression of secondary hyperparathyroidism in the context of hypomagnesemia supported by the high prevalence of hypomagnesemia in COVID-19 (19). Hypomagnesemia suppresses secondary hyperparathyroidism in osteoporotic patients (20) but there is not strong evidence of the impact of hypomagnesemia on calcic metabolism in COVID-19 (12,13).

Parathyroid gland may also be impaired in the inflammatory response during COVID-19 (13). Contrary to our results, Need AG et al. described an association between 25(OH)D deficiency and ionized calcium in young non-COVID-19 patients, particularly when the 25(OH)D level is below 10 nmol/L (21). Under this concentration, the mechanism of secondary hyperparathyroidism is triggered in healthy patients (21).

Thus, COVID-19 is certainly a factor of blunted secondary hyperparathyroidism. This is, to our knowledge, the first study to expose such results in older patients in the context of COVID-19. Furthermore, total and corrected calcium were shown negatively associated with 25(OH)D deficiency, leading to speculate that 25(OH)D interacts not only with ionized calcium but also with calcium-bound proteins. The 1,25(OH)D stimulates the production of calmodulin, a protein known to promote intracellular calcium transport in duodenal cell (22). It may stimulate other proteins involved in calcium metabolism and research should be conducted in this direction.

Moreover, elevated levels of unbound and unsaturated fatty acids have been reported in patients with COVID-19. Unsaturated fatty acids can bind calcium and cause hypocalcemia (23,24). This may explain a modification in [ionized calcium/calcium-bound to proteins] ratio explaining the absence of association between ionized calcium and 25(OH)D deficiency.

The secondary results raise questions about the relevance of calcium correction with the Payne formula in the older population. A better correlation between ionized calcium and total calcium than adjusted calcium and ionized calcium was observed. This result is not described elsewhere to our knowledge. However, some studies suggest that misclassification of calcium status is frequent in the older patients with adjusted calcium (9,10). These results may lead to further study in the older population including analysis of hypomagnesemia.

Limitations should be noted. First, we did not measure 1,25(OH)D, due to costs concern. The analysis of the association between the different types of calcium with the active part of vitamin D could have allowed us to be more precise in understanding the mechanism of secondary hyperparathyroidism in COVID -19. Second, the retrospective nature of this study precludes an analysis of dynamic changes in phospho-calcic metabolism. Third, modification of the ionized calcium assay technique during the study may have affected the reproducibility of the ionized calcium measurement. Fourth, debates on the threshold of 25(OH)D deficiency regularly feed the international literature; we have chosen a 25(OH)D concentration inferior to 50 nmol/L according to OMS recommendation and published studies on the subject (2,13). Fifth, inclusion in this study was mostly in winter and fall despite variabilities in the level of 25(OH)D during the year.

This study highlights the need for a better understanding of phospho-calcic metabolism and offers interesting avenues of research. The perturbations of phospho-calcic metabolism frequently observed in COVID-19 are present in other viral pathologies such as EBOLA virus or H7N9 avian influenza, suggesting a viral pathway of phospho-calcium metabolism abnormalities (11,25).

## CONCLUSION

In conclusion, we found that in the older patients with COVID-19, 25(OH)D deficiency were associated with total and corrected calcium. We described for the first time in older patients the lack of response of secondary hyperparathyroidism to 25(OH)D deficiency in COVID-19. Further studies are therefore needed to better explain the disturbances in phospho-calcium metabolism in both the elderly and COVID population.

## Data Availability

All relevant data are within the manuscript and its Supporting Information files.

## ACKNOWLEDGMENTS

The authors wish to thank the GERIA-COVID study group. GERIA-COVID study group: Cédric Annweiler^1^, Marine Asfar^1^, Mélinda Beaudenon^1^, Jean Barré^1^, Antoine Brangier^1^, Mathieu Corvaisier^1^, Guillaume Duval^1^, Jennifer Gautier^1^, Mialy Guenet^1^, Jocelyne Loison^1^, Frédéric Noublanche^1^, Marie Otekpo^1^, Hélène Rivière^1^, Guillaume Sacco^1^, Romain Simon^1^. Affiliations: 1: Department of Geriatric Medicine, University Hospital, Angers, France.

The authors have listed everyone who contributed significantly to the work in the Acknowledgments section. Permission has been obtained from all persons named in the Acknowledgments section. There was no compensation for this contribution.

## SPONSOR ROLES

None.

## AUTHOR CONTRIBUTIONS

- CA has full access to all of the data in the study, takes responsibility for the data, the analyses and interpretation and has the right to publish any and all data, separate and apart from the attitudes of the sponsors. All authors have read and approved the manuscript.
- Study concept and design: AB, MA, CA.
- Acquisition of data: MA, JG and CA.
- Analysis and interpretation of data: AB, JG and MA.
- Drafting of the manuscript: AB and MA.
- Critical revision of the manuscript for important intellectual content: GS, MA and JG.
- Obtained funding: Not applicable.
- Statistical expertise: JG.
- Administrative, technical, or material support: CA.
- Study supervision: CA.

## DATA AVAILABILITY

Patient level data are freely available from the corresponding author at. There is no personal identification risk within this anonymized raw data, which is available after notification and authorization of the competent authorities.

